# A Novel Question-Answering Framework for Automated Abstract Screening Using Large Language Models

**DOI:** 10.1101/2023.12.17.23300102

**Authors:** Opeoluwa Akinseloyin, Xiaorui Jiang, Vasile Palade

## Abstract

**Objective:** This paper aims to address the challenges in abstract screening within Systematic Reviews (SR) by leveraging the zero-shot capabilities of large language models (LLMs).

**Methods:** We employ LLM to prioritise candidate studies by aligning abstracts with the selection criteria outlined in an SR protocol. Abstract screening was transformed into a novel question-answering (QA) framework, treating each selection criterion as a question addressed by LLM. The framework involves breaking down the selection criteria into multiple questions, properly prompting LLM to answer each question, scoring and re-ranking each answer, and combining the responses to make nuanced inclusion or exclusion decisions.

**Results:** Large-scale validation was performed on the benchmark of CLEF eHealth 2019 Task 2: Technology-Assisted Reviews in Empirical Medicine. Focusing on GPT-3.5 as a case study, the proposed QA framework consistently exhibited a clear advantage over traditional information retrieval approaches and bespoke BERT-family models that were fine-tuned for prioritising candidate studies (i.e., from the BERT to PubMedBERT) across 31 datasets of four categories of SRs, underscoring their high potential in facilitating abstract screening.

**Conclusion:** Investigation justified the indispensable value of leveraging selection criteria to improve the performance of automated abstract screening. LLMs demonstrated proficiency in prioritising candidate studies for abstract screening using the proposed QA framework. Significant performance improvements were obtained by re-ranking answers using the semantic alignment between abstracts and selection criteria. This further highlighted the pertinence of utilizing selection criteria to enhance abstract screening.

## Introduction

A Systematic Review (SR) in medical research is the highest form of knowledge synthesis of all available medical evidence from relevant publications on a specific topic. SR follows a principled pipeline, including candidate study retrieval, primary study selection, quality assessment, data extraction, data synthesis, meta-analysis, and reporting [1]. Because of its thoroughness and reliability, SR underpins evidence-based medicine [2]. It shapes medical research and practice by informing researchers of the state-of-the-art knowledge and knowledge gaps as well as health practitioners and policymakers of the best clinical practice [3].

SR also faces tremendous challenges at each step. For instance, it is time-consuming, expensive and resource-intensive to select primary studies, a.k.a. *abstract screening*, due to the massive volume of retrieved candidate studies, often at tens of thousands [4, 5]. It is further worsened by involving multiple human annotators, which is required to reduce bias and disparities [6]. This compound complexity calls for innovative solutions to automate or semi-automate abstract screening [1] to minimize the time delays and costs of this manual screening task [7], which is the focus of the current paper. Figure 1 shows an example of abstract screening, where the abstract of an included study is matched against the selection criteria defined in the SR protocol.

**Fig. 1:**
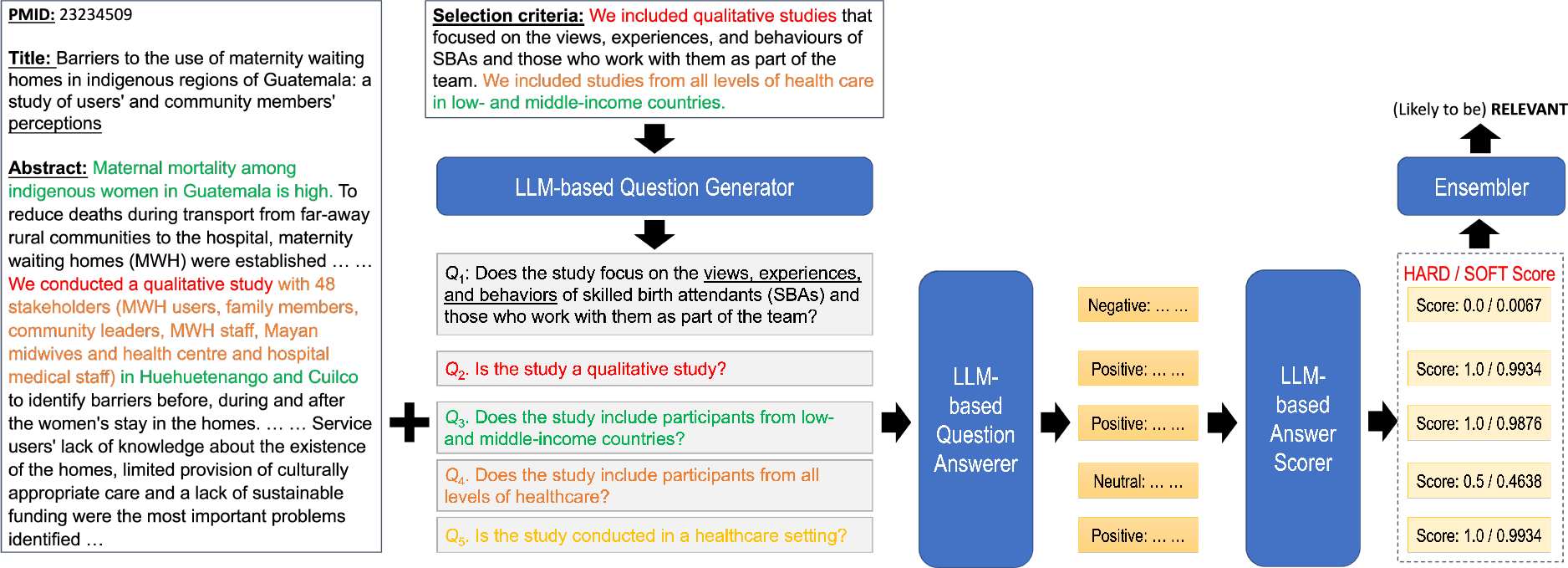
Illustration of LLM-assisted automated abstract screening.

Machine learning has been the focus of research in automating abstract screening [1, 7, 8]. Firstly, a small set of studies is selected for human annotation, and then a classifier is trained. Typically, active learning is adopted to improve the classifier iteratively. Obviously, the quality of the initial annotations plays an important role. However, choosing initial annotations is a problem of zero-shot setting and has not been explored at all. Another disadvantage is that this approach is not generalisable, and each SR topic requires training a bespoke classifier from scratch.

An alternative perspective was to treat abstract screening as a ranking problem a.k.a. *reference prioritisation* [7], incorporating approaches from the information retrieval (IR) community [9, 10, 11, 12, 13, 14, 15, 16]. One advantage of this approach is that it can utilise additional information about an SR converted into queries to enhance screening performance. Such information could be review title [9, 10], original Boolean queries (for candidate study retrieval) [17], research objectives [18, 16], or a set of seed studies [15, 19]. Another advantage is the possibility of training a cross-topic ranker to generalise to diverse SR topics.

The above analysis motivated us to explore the emerging capabilities of Large Language Models (LLMs), particularly GPT-3.5, to facilitate abstract screening. LLMs, with their robust zero-shot capabilities [20], offer the potential to act as AI-based reviewers, streamlining the abstract screening process by either replacing at least one human reviewer or suggesting an initial set of abstracts for human verification and classifier training, both significantly reducing the workload for human reviewers .

In addition, we witness a severe lack of study about using selection criteria in automated abstract screening. Indeed, the selection criteria set up the grounds for human reviewers’ decision-making. Unfortunately, only a few studies initiated similar attempts [21, 22, 23], but neither the effectiveness of their methods nor the comprehensiveness of their experiments could provide convincing conclusions about the feasibility of LLMs in this task. The current paper presents a pioneering LLM-based framework for facilitating automated abstract screening to fill this gap.

Our contributions can be summarised in three folds. (1) We proposed the first comprehensive LLM-assisted question-answering framework to facilitate automated abstract screening in a zero-shot setting. (2) We developed the first generalisable approach to utilising selection criteria to enhance abstract screening efficiency. (3) Our study marks the first comprehensive exploration of leveraging LLMs for reference prioritisation in abstract screening, utilizing a well-known benchmark dataset to showcase the method’s high potential.

## Background Study

### Automation in Abstract Screening

Efforts to automate systematic reviews using machine learning have surged recently. Kitchenham and Charters’ presented a good survey of such attempts in software engineering [24]. In evidence-based medicine, Cohen et al. was the seminal work of abstract screening using machine learning [25], while Marshall and Wallace advocated active learning techniques for abstract screening [26]. Examples like RobotReviewer [27, 28] and TrialStreamer [29] showcased the power of integrating AI into the review process, with RobotReviewer claiming to reach accuracy comparable to human reviewers. Despite the progress, challenges persist, including labour-intensive labelling and the risk of overlooking relevant studies [30]. Acknowledging the limitation of full automation, tools like Rayyan and Abstracker leverage natural language processing (NLP) algorithms to partially automate article screening [31].

### Machine Learning for Abstract Screening

The biggest challenge is handling large document volumes, particularly in non-randomized controlled trials lacking database filters [32]. For instance, EPPI-Centre reviews often screen over 20,000 documents, necessitating more efficient approaches [33]. Efforts include refining search queries, balancing precision and recall, and leveraging resource-efficient recall-maximizing models with NLP [34].

The initial approach involves training a classifier to make explicit include/exclude decisions [25, 34, 35, 36, 37, 38, 39]. Many classifiers using this approach inherently generate a confidence score indicating the likelihood of inclusion or exclusion (similar to the ranking in the second approach). Generally, this approach requires a labelled dataset for training, hindering the assessment of work reduction until after manual screening. Research within this paradigm primarily focuses on enhancing feature extraction methods [25, 37] and refining classifiers [38]. Van Dinter et al. [8] analyzed 41 studies in medicine and software engineering, revealing Support Vector Machines and Bayesian Networks as standard models and Bag of Words and TF-IDF as prevalent natural language processing techniques. Despite advancements, a dearth of deep neural network models explicitly designed for the systematic review screening phase is noted. The most prominent challenges include handling extreme data imbalance favouring (at least close to) total recall of relevant studies.

### Ranking Approaches to Abstract Screening

The second approach entails utilizing a ranking or prioritisation system [9, 10, 11, 12, 13, 14, 15, 16, 33, 40]. This approach might necessitate manual screening by a reviewer until a specified criterion is met. This approach can also reduce the number of items needed to be screened when a cut-off criterion is properly established [33, 40, 41]. In addition to reducing the number needed to screen, other benefits of this approach include enhancing reviewers’ understanding of inclusion criteria early in the process, starting full-text retrieval sooner, and potentially speeding up the screening process as confidence in relevance grows [7]. This prioritisation approach also aids review updates, enabling quicker assimilation of current developments. Various studies reported benefits from prioritisation for workflow improvement, emphasizing efficiency beyond reducing title and abstract screening workload [42, 43].

### Active learning in Abstract Screening

It’s crucial to note that the last approach, active learning, aligns with both strategies above [34, 33, 44]. This involves an iterative process to enhance machine predictions by interacting with reviewers. The machine learns from an initial set of include/exclude decisions human reviewers provide. Reviewers then judge a few new samples, and the machine adapts its decision rule based on this feedback. This iterative process continues until a specified stopping criterion is met. While the classifier makes final decisions for unscreened items, human screeners retain control over the training process and the point at which manual screening concludes. Wallace et al. implement active learning-based article screening using Support Vector Machines [34]. Notable tools include Abstrackr [36] and ASReview [45]. Various active learning strategies existed [7]. For instance, Marshall and Wallace [26] proposed a variant based on certainty, continuously training the classifier on manually screened articles and reordering unseen articles based on predicted relevance.

### Large Language Models for Abstract Screening

Recent advancements in LLMs, notably demonstrated by ChatGPT (GPT-3.5 or 4.0), have brought about a revolutionary paradigm shift across disciplines [46, 47]. LLMs have shown impressive generalisability across diverse domains and strong zero-/few-shot reasoning capabilities [46, 48]. Leveraging LLMs holds promise for SRs, which, however, remains underexplored [7, 8]. This gap underscores the need for a comprehensive investigation into LLMs’ potential in automating SRs, e.g., abstract screening in the current paper.

There are some initial attempts to evaluate ChatGPTs in automated SR, such as automating search queries [49]. Alshami et al. [50] leveraged ChatGPT to automate the SR pipeline, yet their methodology diverges from conventional abstract screening practices, rendering it distinct from traditional approaches. The application of ChatGPT in abstract screening has been scarcely explored. Only two studies tried to address it [51, 52]. However, these studies failed to achieve a high recall rate (preferably total or close to total recall, say 95% in most studies), a critical factor for practical applicability. In addition, these studies performed limited empirical studies on a small number of in-house datasets that were neither public nor common in the research community, making it even harder to do reliable effectiveness evaluation. In contrast, our paper introduces a novel approach by applying LLMs like GPT-3.5 for reference prioritisation across a substantially larger and well-recognised benchmark [53], significantly enhancing the method’s applicability and effectiveness in systematic reviews.

## Materials and Methods

### Overview

Our framework utilizes LLMs’ remarkable zero-shot learning ability to assess if a candidate study’s abstract aligns with the SR protocol’s selection criteria. These criteria outline aspects of the selected studies. Figure 1 illustrates the idea using an example. The inclusion criteria contain four parts (in different colours). Each can be answered by matching the relevant information in the abstract (in corresponding colours). For instance, the selection criterion “We included qualitative studies” is answered by the text evidence “We conducted a qualitative study” (both in red). Theoretically, all inclusion criteria should be met for the study to be included in the SR.

The current paper frames automated abstract screening as a question-answering (QA) task and proposes to use LLMs like GPT-3.5 to solve it. LLMs have showcased impressive question-answering abilities across diverse domains and tasks, including encoding clinical knowledge and achieving success in medical licensing exams [54, 55, 56, 57]. Initially, we experimented with condensing the entire selection criteria into a single, comprehensive question, as depicted in Figure 2a. This method quickly showed its limitations, as LLMs are notably more adept at addressing well-defined, focused questions. Recognizing the critical need for high recall—95% or higher—to preserve the comprehensive integrity of the SR, we shifted our strategy to treating each selection criterion as a question to be addressed using LLMs. For example, the four selection criteria in Figure 1 are converted into questions *Q* 1 to *Q* 4 (by the LM-based Query Generator component in Figure 1). Subsequent prompts are directed to LLMs to get individual answers (from the LM-based Question Answerer), which are then combined into the final decision (through the Answer Scorer and Ensembler). Note that, in this example, *Q* 5 is basically a useless, redundant question because we hard-coded GPT-3.5 to generate five questions (also refer to the “Question Generation” section).

**Fig. 2:**
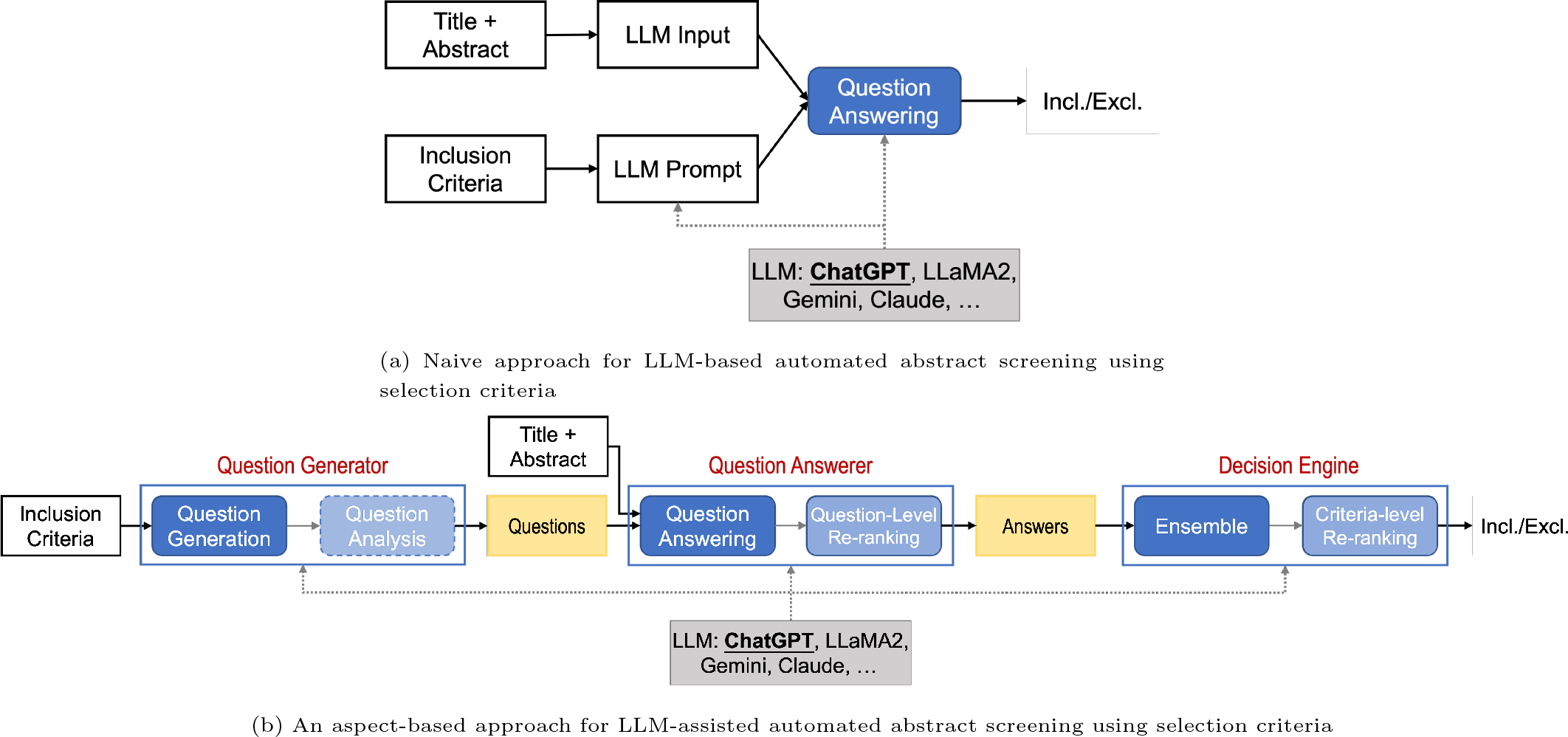
Methodological framework for LLM-assisted automation screening.

Figure 2b formally defines our proposed QA framework for abstract screening. The following subsections will detail each component. We begin with a Question Generator to convert the selection criteria into a set of questions. Question generation is done by LLM. Optionally, question analysis may be done to analyse the logic between questions for the purpose of correctly combining question answers. See the “Quality of Question Generation” for details. Subsequently, each question is addressed by a trained Question Answerer to determine if the corresponding selection criterion is met. Question answering is also done by LLM. Each question answer is converted into a numeric score, and question-level re-ranking will be applied to improve answer scores. While the current paper tried some primitive but effective ways for answer re-ranking, the framework is flexible enough to plug in more advanced re-ranking methods. Finally, the Decision Engine ensembles the answers to all questions (by aggregating answer scores), followed by an optional criteria-level re-ranking step to calibrate the final score for decision-making.

### Question Generation

A substantial body of research exists on automated question generation from natural language text [58]. These methods often rely on manually crafted rules or a trained model, typically a fine-tuned pre-trained language model. While these question generation models have demonstrated utility in domain-specific tasks, such as generating questions about product descriptions for matching purchase inquiries[59] or creating questions about academic materials to assess learning outcomes [60], generalizing them to the vast diversity of SR topics presents challenges. Therefore, we entrust the question generation task to LLMs, here GPT-3.5.

### Prompt Design

Our prompt development process adhered to OpenAI’s guidelines [61]. We went further and established a persona for the LLM, instructing it: “‘You are a researcher screening titles and abstracts of scientific papers for the systematic review.” (Fig. 3a). This persona was crucial in ensuring the LLM’s responses were accurately contextualised so that they properly aligned with the task objective. Delimiters were used to separate different sections of the prompt, enhancing clarity and coherence. A naive approach to question generation is to prompt the LLM to generate questions from the selection criteria paragraph. However, the early evaluation revealed that this uncontrolled method often generated redundant, duplicate or sometimes trivial questions. To enhance the quality of generated questions, we constrained GPT-3.5 to produce no more than *K* questions. Based on an analysis of the lengths of selection criteria in our datasets, *K* = 5 proved sufficient for most SRs. This explicitly instructed the LLM to avoid redundancy. Figure 3a depicts the utilized prompt, and an example is shown in Figure 1. Appendix A in the online supplementary material contains all the questions generated from the corresponding selection criteria for all SRs in the benchmark we evaluated. Each sentence in the selection criteria often aligns with a distinct criterion. In rare cases with more than 5 sentences, GPT-3.5 intelligently combined two sentences into one question. In addition to diversity, we also found better quality of the questions (See the “Quality of Question Generation” section for the quantitative evaluation).

**Fig. 3:**
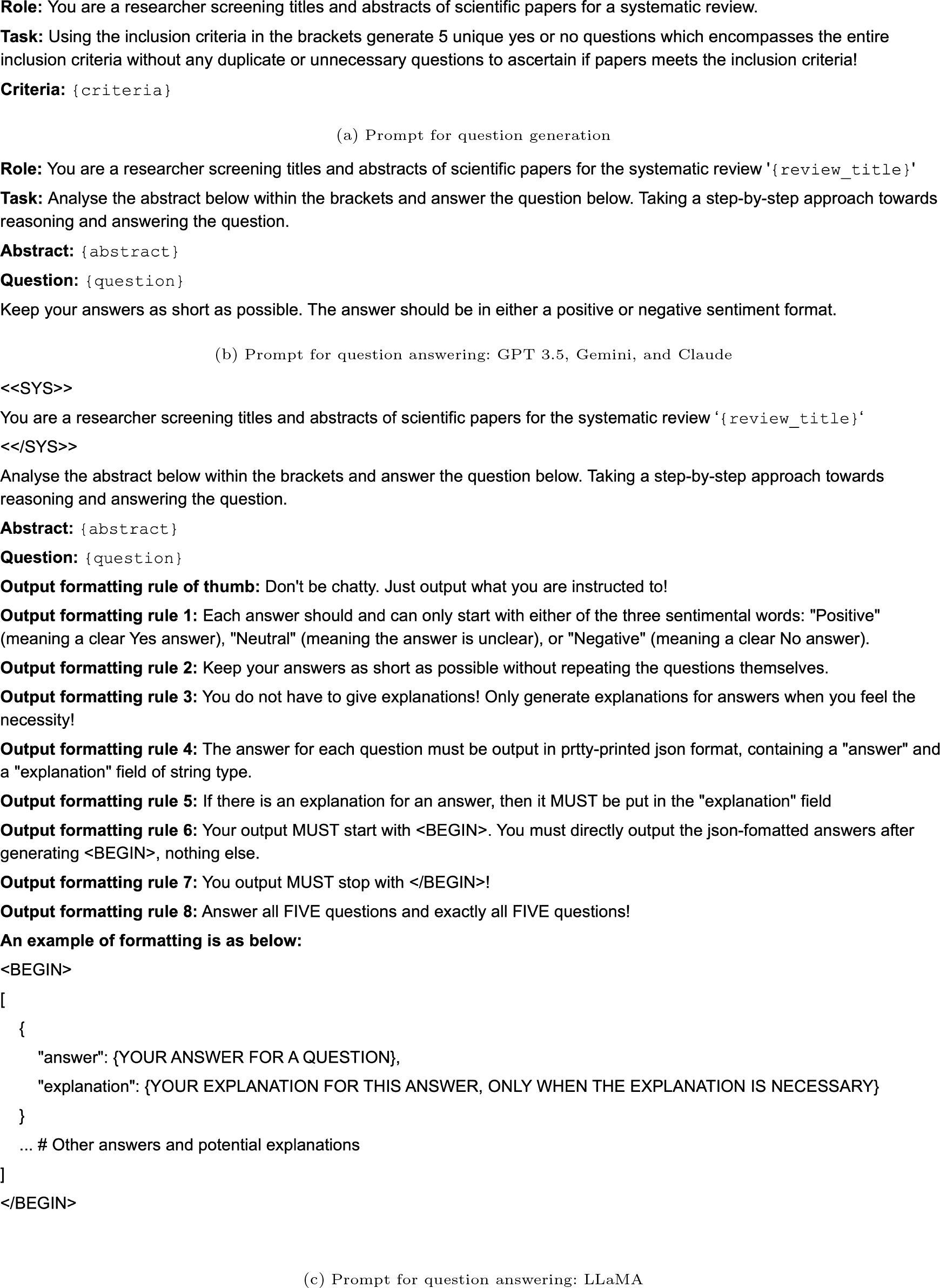
Prompt design for LLM-assisted automated abstract screening

### Question Answering

The Question Answerer evaluates the relevance of each abstract to every selection criterion, formulated as Yes/No questions. Initial tests showed that LLMs generally explain their answers. To quantify these explanations, we incorporated sentiment analysis to score the responses (see the “Answer Scoring” section). We instructed LLMs (Figure 3b) to reply either a “Positive”, “Negative”, or “Neutral” answer:

*•* **Positive:** The abstract explicitly addresses the question, offering information that aligns with the criteria posed by the question.
*•* **Neutral:** The information in the abstract is inadequate or too ambiguous for LLMs to derive a confirmatory answer.
*•* **Negative:** A clear NO answer to the question, indicating irrelevance to the specified criteria.

### Prompt Design

The same prompt was used by most LLMs (Figure 3b). It was meticulously designed using three randomly selected topics from the Intervention training set of the TAR2019 benchmark. This approach prevented information leakage during prompt development, ensuring methodological integrity. The same persona was used for both question generation and answering. The only exception happened with LLaMA. We found LLaMA was unable to output the answers (and explanations) in a consistent style for post-processing. Thus, we tested a number of output formatting rules to (i) “force” LLaMA to answer only what is needed to be answered, and (ii) to “guide” LLaMA to format its answers (and explanations) in an easy-to-process JSON format, as Figure 3c shows.

### Answer Scoring

LLMs often generate an explanation for their answer. We converted answers into numeric representations using BART as a zero-shot sentiment scorer [62]. The problem definition is formlated as follows. Suppose the set of candidate studies are 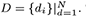. The system review protocol defines a set of selection criteria (questions) 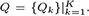. Given each candidate study *d_i_*, the set of answers are 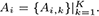. Given a candidate study *d_i_* and each question answer *A_i,k_*, we defined two methods, namely the Hard Answer and Soft Answer, to assign the answer score, denoted as *score*(*d_i_, A_i,k_*), using BART, which reflects the likelihood of an abstract meeting a selection criterion.

### Hard Answer

In the Hard Answer method, BART classifies LLMs’ responses into three distinct categories: Positive, Neutral, and Negative, and accordingly, we assign a fixed score 1, 0.5, and 0 to each category, denoted by *score*(*d_i_, A_i,k_*). Thus,

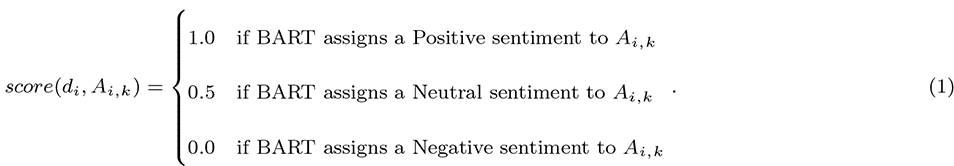

### Soft Answer

In the Soft Answer method, each answer is scored by the probability of its sentiment being positive, which is calculated by BART. Thus,

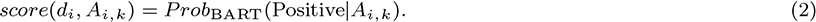

### Question-level Re-ranking

To enhance screening further, one significant contribution of the current paper involves re-ranking candidate studies based on how well abstracts are semantically aligned with the selection criteria. For each Yes/No question, the cosine similarity between the question (selection criterion) and the abstract is computed based on their text embeddings encoded by an LLM [63], and averaged with the original answer score, producing *K* re-ranked scores, defined by

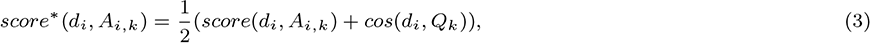

where *cos*(*d_i_, Q_k_*) is the cosine similarity between (the abstract of) a candidate study *d_i_* and one selection criterion question *Q_k_*.

### Decision Engine

#### Ensemble

Given a candidate study *d_i_*, the answer scores for each selection criterion are “averaged” as the score for the candidate study, denoted by *score*(*d_i_*):

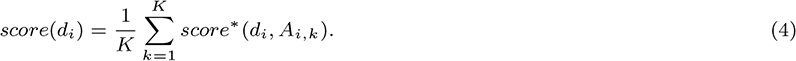

#### Criteria-level Re-ranking

criteria-level re-ranking is done by averaging the score for candidate study *d_i_* with the cosine similarity between its abstract and the selection criteria *Q*. Note that here, we use the original selection criteria paragraph in the review protocol for calculation instead of the questions generated from it. Thus, the re-ranked score for *d_i_*, denoted by *score^∗^*(*d_i_*), is calculated by

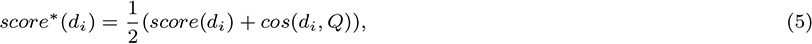

where *cos*(*d_i_, Q*) is the cosine similarity between the candidate study and the complete selection criteria. Candidate studies are prioritised in descending order of this final score *score^∗^*(*d_i_*).

### Experimental Setup

#### Dataset and Evaluation Metrics

This study utilized the widely-used benchmark for evaluation: CLEF eHealth 2019 Task 2: Technology-Assisted Reviews in Empirical Medicine (TAR2019) [53]. This benchmark provides valuable insights into the prevailing scientific consensus on various topics, making it a suitable resource for evaluating re-ranking methodologies in systematic reviews [64]. We employed the TAR2019 test set comprising 31 SRs grouped into four topic categories: 20 about clinical intervention trials (Intervention), eight about diagnostic technology assessment (DTA), 2 for qualitative studies (Qualitative), and 1 for Prognosis. Appendix B in the online supplementary material contains detailed statistics of the SRs in each category. We refrained from using the training set provided by TAR2019, aiming to highlight the effectiveness of our zero-shot methodology that eliminates the need for prior training [65, 23]. Selection criteria for each SR are included in TAR2019. Seven evaluation metrics were employed, including the rank position of the last relevant document (*L Rel*), Mean Average Precision (*MAP*), Recall at *k* % (*R*@*k*%, *k* = 5, 10, 20, 30), and Work Saved Over Sampling (WSS) at a *r* % recall level (*WSS*@*r*%, *r* = 95, 100). Notably, *WSS*@*r*% measures the screening workload saved by halting the examination process once *r* % of relevant documents are identified, compared to screening the entire document set [25]. For each SR, we calculated the performance metrics. They were averaged over all the SRs in each category.

#### Baseline Models

We compared our methods against a number of zero-short baselines, including Sheffield University’s submission [17] to TAR2019 and the six BERT-based ranking models by Wang et al. [23]. To comprehensively assess performance, we implemented two IR baselines of our own. One is cosine similarity between selection criteria and abstract based on GPT embeddings [63], named GPT Cos Sim Criteria. The other is the classical IR approach BM25 [66], using selection criteria as a query to rank candidate studies. The baselines are summarised below:

*•* sheffield-baseline: The zero-shot IR baseline submitted by the University of Sheffield to TAR2019.
*•* QLM: Another IR baseline in the Query Likelihood Model. See Wang et al. [23] for details.
*•* BERT/BERT-M/BioBERT/BlueBERT/PubMedBERT: Use BERT-family models as neural rankers, including BERT [67], BERT-M (BERT fine-tuned on the MS MARCO machine reading comprehension dataset) [68], and BERT models tailed to biomedical domains, such as BioBERT [69], BlueBERT [70] and PubMedBERT [71].

#### Large Language Models

While we focus on GPT-3.5 (more precisely “gpt-3.5-turbo-16K”) as a case study to demonstrate the effectiveness of the proposed framework, three additional successful mainstream LLMs were also employed on the more challenging *DTA* subset on which GPT-3.5 underperformed: LLaMA 2 (“llama-2-70b-chat-hf”), Gemini (“gemini-pro”), and Claude 3 (“claude-3-haiku-20240307”). The other rationale for choosing these LLMs for question answering was model “comparability”. LLaMA 2 was seen as a comparable LLM to GPT-3.5. While both Gemini and Claude are closed-source LLMs, making direct comparison difficult, we used API cost as a proxy for their capabilities in answering selection criterion questions and decided to test “gemini-pro” and “claude-3-haiku-20240307”. Besides, Claude is a more recent leading proprietary LLM, so we also expect to see a positive impact on the quality of question answering. For all models, we used a temperature setting of 0.2. In rare cases when LLMs failed to give answers to all the questions after a number of tries, we prompted LLMs to answer each question one by one. For answer re-ranking, becasue LLaMA and Claude do not provide their own text embedding models, we focused on GPT embedding (“text-embedding-ada-002”) in our main results (the “Effectiveness of the Question-Answering Framework” section), and also covered Gemini embedding (“text-embedding-004”) in LLM comparisons (see the “Comparing Large Language Models” section).

## Results and Discussions

### Effectiveness of Using Selection Criteria

Our first experiments aimed to evaluate the effectiveness of selection criteria for abstract screening. For comparison, we also used review title and search queries, both included in the TAR2019 benchmark, for prioritising candidate studies in a similar way. As the “Large Language Models” section explains, GPT embedding was used in this experiment.

The initial results in Table 1 demonstrate that selection criteria contain indispensable information for abstract screening. For example, on the *Intervention* and *DTA* datasets, the top 5% ranked results covered 40.1% and 47.7% included studies, respectively, while the top 30% covered 79.7% and 85.1% included studies, respectively. This means selection criteria help to push a significant number of included studies to the front of the ranking lists, demonstrating its potential for prioritising candidate studies. The *WSS*@95% values mean that at most 52.2% and 60% manual screening time can be saved to achieve a 95% recall of included studies. On *Qualitative* datasets, *R*@10% was 47.8%, still very high, but the *WSS* values were not as good as on *Intervention* and *DTA*. *R*@*k*% started to stagnate when *k* grew (e.g., beyond 30%).

**Table 1.** The results of using three types of queries to match abstracts of candidate studies: selection criteria, review titles, and search queries.

| Dataset | Models | L_Rel | MAP | R@5% | R@10% | R@20% | R@30% | R@50% | WSS@95% | WSS@100% |
| --- | --- | --- | --- | --- | --- | --- | --- | --- | --- | --- |
| Intervention | Review Title | 927 | 0.344 | 0.441 | 0.596 | 0.758 | 0.836 | 0.929 | 0.560 | 0.519 |
|  | Search Query | 1248 | 0.200 | 0.241 | 0.424 | 0.602 | 0.725 | 0.889 | 0.449 | 0.416 |
|  | Selection Criteria | 920 | 0.315 | 0.401 | 0.544 | 0.722 | 0.797 | 0.797 | 0.522 | 0.499 |
| DTA | Review Title | 1379 | 0.170 | 0.283 | 0.497 | 0.678 | 0.807 | 0.926 | 0.525 | 0.446 |
|  | Search Query | 1593 | 0.203 | 0.228 | 0.412 | 0.658 | 0.781 | 0.909 | 0.473 | 0.417 |
|  | Selection Criteria | 1154 | 0.271 | 0.477 | 0.628 | 0.782 | 0.851 | 0.851 | 0.600 | 0.513 |
| Qualitative | Review Title | 1992 | 0.113 | 0.200 | 0.278 | 0.609 | 0.650 | 0.950 | 0.506 | 0.406 |
|  | Search Query | 2221 | 0.046 | 0.105 | 0.173 | 0.255 | 0.591 | 0.923 | 0.369 | 0.329 |
|  | Selection Criteria | 2256 | 0.082 | 0.173 | 0.478 | 0.559 | 0.618 | 0.618 | 0.303 | 0.289 |
| Prognosis | Review Title | 2983 | 0.136 | 0.147 | 0.258 | 0.495 | 0.668 | 0.837 | 0.247 | 0.106 |
|  | Search Query | 3017 | 0.200 | 0.237 | 0.400 | 0.605 | 0.768 | 0.900 | 0.311 | 0.096 |
|  | Selection Criteria | 3160 | 0.178 | 0.200 | 0.305 | 0.495 | 0.647 | 0.832 | 0.239 | 0.053 |

It is also observed that selection criteria significantly outperformed search query on the *Intervention* and *DTA* datasets, according to most performance metrics except *R*@50%. Overall, review title seems to be most valuable for screening candidate studies, so it is also part of our prompt design (Figure 3). Search Query was less effective. Indeed, the impreciseness of search queries resulted in a large number of studies being screened. The only exception was *Prognosis*, but the fact that the *Prognosis* category contains just one SR does not allow us to overturn the afore-made overall statements.

Acknowledging such contextual variability inherent in the initial results is crucial. The nature of the dataset appeared to play a substantial role in the effectiveness of selection criteria. For instance, datasets with more specific interventions or defined methodologies, like *DTA*, may inherently align more closely with selection criteria, enhancing the performance of this query type in prioritisation. In conclusion, the above evidence **advocates for the integration of selection criteria in abstract screening** automation tools.

### Effectiveness of the Question-Answering Framework

In this subsection, we summarise the main findings based on experiments using GPT-3.5 for both question answering and answer re-ranking. Table 2 compares our QA-based approaches (i.e., the GPT QA Soft Both ReRank and GPT QA Hard Both ReRank two rows) against other zero-shot baselines. “Both ReRank” means both re-raking methods were used. The GPT Cos Sim Criteria rows are copied from Table 1. Consistent with the baselines, we reported the average performances of each metric for each SR topic category. Appendix C contains the results for each individual SR. On four topic categories, our approaches outperformed almost all baselines across a multitude of metrics. Specifically, our approaches scored substantially better in *L Rel* (the lower, the better) and *MAP* (the higher, the better), indicating the potential for pruning irrelevant studies more efficiently (low *L Rel*) and accurately (high *MAP*). Moreover, *WSS*@95% and *WSS*@100% both significantly outperformed the baselines, except Cosine Similarity Criteria on *DTA*, indicating the potential for saving more work from human reviewers who seek a total or close-to-total recall of included studies.

**Table 2.**
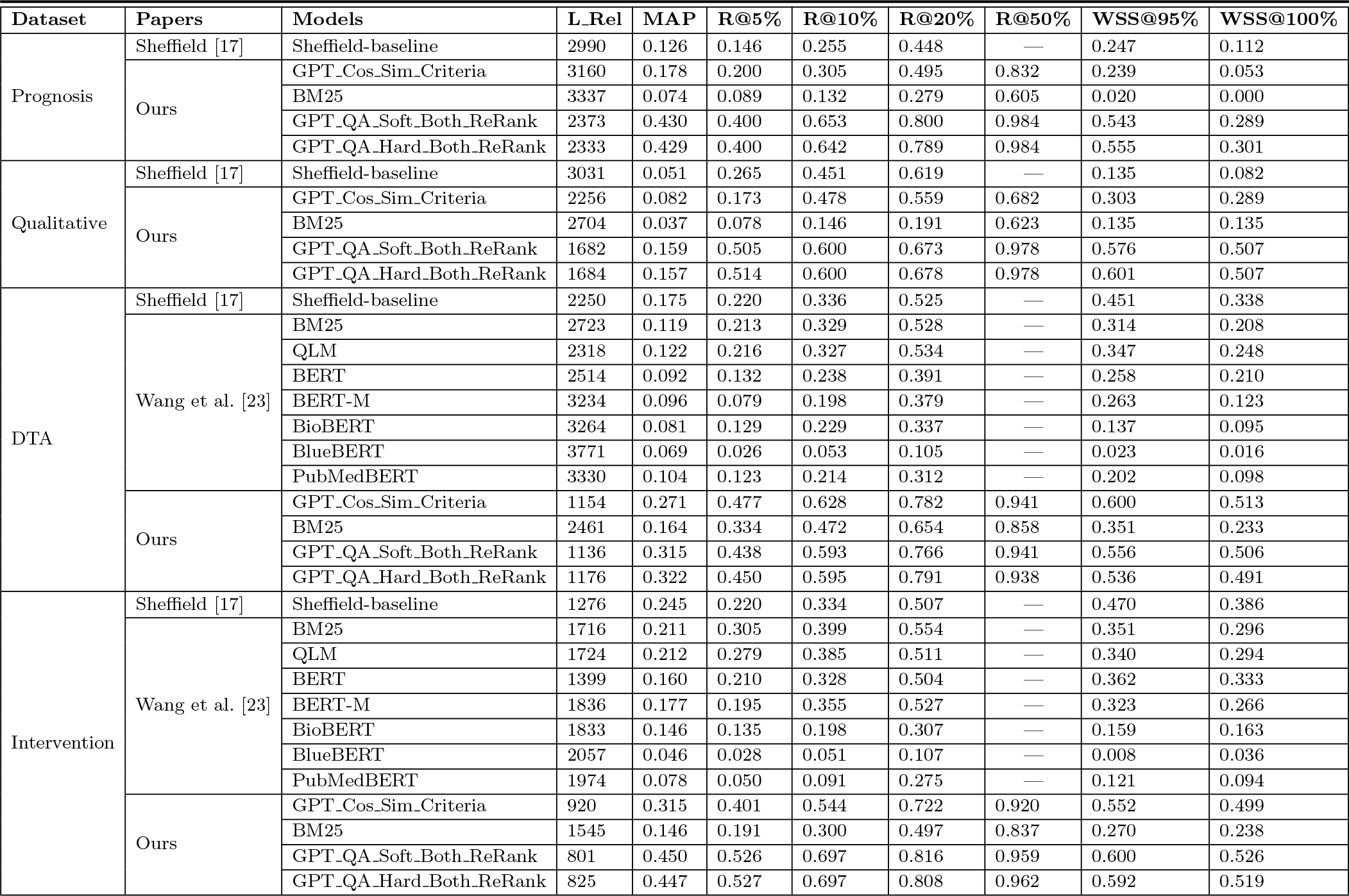
Overview and comparisons of model performances on four systematic review categories. Note that the baselines did not report *R*@50%.

| Dataset | Papers | Models | *L*_Rel | MAP | R@5% | R@10% | R@20% | R@50% | WSS@95% | WSS@100% |
| --- | --- | --- | --- | --- | --- | --- | --- | --- | --- | --- |
| Prognosis | Sheffield [17] | Sheffield-baseline | 2990 | 0.126 | 0.146 | 0.255 | 0.448 | — | 0.247 | 0.112 |
|  | Ours | GPT_Cos_Sim_Criteria | 3160 | 0.178 | 0.200 | 0.305 | 0.495 | 0.832 | 0.239 | 0.053 |
|  |  | BM25 | 3337 | 0.074 | 0.089 | 0.132 | 0.279 | 0.605 | 0.020 | 0.000 |
|  |  | GPT_QA_Soft_Both_ReRank | 2373 | 0.430 | 0.400 | 0.653 | 0.800 | 0.984 | 0.543 | 0.289 |
|  |  | GPT_QA_Hard_Both_ReRank | 2333 | 0.429 | 0.400 | 0.642 | 0.789 | 0.984 | 0.555 | 0.301 |
| Qualitative | Sheffield [17] | Sheffield-baseline | 3031 | 0.051 | 0.265 | 0.451 | 0.619 | — | 0.135 | 0.082 |
|  | Ours | GPT_Cos_Sim_Criteria | 2256 | 0.082 | 0.173 | 0.478 | 0.559 | 0.682 | 0.303 | 0.289 |
|  |  | BM25 | 2704 | 0.037 | 0.078 | 0.146 | 0.191 | 0.623 | 0.135 | 0.135 |
|  |  | GPT_QA_Soft_Both_ReRank | 1682 | 0.159 | 0.505 | 0.600 | 0.673 | 0.978 | 0.576 | 0.507 |
|  |  | GPT_QA_Hard_Both_ReRank | 1684 | 0.157 | 0.514 | 0.600 | 0.678 | 0.978 | 0.601 | 0.507 |
| DTA | Sheffield [17] | Sheffield-baseline | 2250 | 0.175 | 0.220 | 0.336 | 0.525 | — | 0.451 | 0.338 |
|  | Wang et al. [23] | BM25 | 2723 | 0.119 | 0.213 | 0.329 | 0.528 | — | 0.314 | 0.208 |
|  |  | QLM | 2318 | 0.122 | 0.216 | 0.327 | 0.534 | — | 0.347 | 0.248 |
|  |  | BERT | 2514 | 0.092 | 0.132 | 0.238 | 0.391 | — | 0.258 | 0.210 |
|  |  | BERT-M | 3234 | 0.096 | 0.079 | 0.198 | 0.379 | — | 0.263 | 0.123 |
|  |  | BioBERT | 3264 | 0.081 | 0.129 | 0.229 | 0.337 | — | 0.137 | 0.095 |
|  |  | BlueBERT | 3771 | 0.069 | 0.026 | 0.053 | 0.105 | — | 0.023 | 0.016 |
|  |  | PubMedBERT | 3330 | 0.104 | 0.123 | 0.214 | 0.312 | — | 0.202 | 0.098 |
|  | Ours | GPT_Cos_Sim_Criteria | 1154 | 0.271 | 0.477 | 0.628 | 0.782 | 0.941 | 0.600 | 0.513 |
|  |  | BM25 | 2461 | 0.164 | 0.334 | 0.472 | 0.654 | 0.858 | 0.351 | 0.233 |
|  |  | GPT_QA_Soft_Both_ReRank | 1136 | 0.315 | 0.438 | 0.593 | 0.766 | 0.941 | 0.556 | 0.506 |
|  |  | GPT_QA_Hard_Both_ReRank | 1176 | 0.322 | 0.450 | 0.595 | 0.791 | 0.938 | 0.536 | 0.491 |
| Intervention | Sheffield [17] | Sheffield-baseline | 1276 | 0.245 | 0.220 | 0.334 | 0.507 | — | 0.470 | 0.386 |
|  | Wang et al. [23] | BM25 | 1716 | 0.211 | 0.305 | 0.399 | 0.554 | — | 0.351 | 0.296 |
|  |  | QLM | 1724 | 0.212 | 0.279 | 0.385 | 0.511 | — | 0.340 | 0.294 |
|  |  | BERT | 1399 | 0.160 | 0.210 | 0.328 | 0.504 | — | 0.362 | 0.333 |
|  |  | BERT-M | 1836 | 0.177 | 0.195 | 0.355 | 0.527 | — | 0.323 | 0.266 |
|  |  | BioBERT | 1833 | 0.146 | 0.135 | 0.198 | 0.307 | — | 0.159 | 0.163 |
|  |  | BlueBERT | 2057 | 0.046 | 0.028 | 0.051 | 0.107 | — | 0.008 | 0.036 |
|  |  | PubMedBERT | 1974 | 0.078 | 0.050 | 0.091 | 0.275 | — | 0.121 | 0.094 |
|  | Ours | GPT_Cos_Sim_Criteria | 920 | 0.315 | 0.401 | 0.544 | 0.722 | 0.920 | 0.552 | 0.499 |
|  |  | BM25 | 1545 | 0.146 | 0.191 | 0.300 | 0.497 | 0.837 | 0.270 | 0.238 |
|  |  | GPT_QA_Soft_Both_ReRank | 801 | 0.450 | 0.526 | 0.697 | 0.816 | 0.959 | 0.600 | 0.526 |
|  |  | GPT_QA_Hard_Both_ReRank | 825 | 0.447 | 0.527 | 0.697 | 0.808 | 0.962 | 0.592 | 0.519 |

The recall values at various percentages (*R*@5%, *R*@10%, *R*@20%) were particularly encouraging. Notably, *R*@5% shows that the top-5% ranked results covered more than 40% (40%-52.7%) of included studies (e.g., by GPT QA Hard Both ReRank 52.7% on *Intervention*, 45% on *DTA*, 51.4% on *Qualitative* and 40% on *Prognosis*). When increased to top-10%, roughly 60-70% of included studies were returned (e.g., 69.7%, 59.5%, 60% and 64.2%, respectively). This result is of great significance. On the one hand, human reviewers could obtain 60% of included studies by verifying only 10% of candidate studies, five times saving compared to random sampling. This allows for **creating a large-enough annotated dataset more efficiently** to train a robust classifier for automated screening [25]. On the other hand, *R*@50% achieved 95% recall on average. This roughly means that the 50% candidate studies ranked in the second half can be safely discarded, which essentially means treating **LLM as the second reviewer** or be delegated to automated screening, both resulting in a roughly 50% saving in the manual annotation. Future work is needed to validate the proposal of using LLM as the second reviewer on more SRs of a wider range of topics.

In summary, our QA-based approach exhibited a clear advantage over traditional models and bespoke BERT-family models that were fine-tuned for ranking tasks (i.e., the BERT to PubMedBERT rows in Table 2). They showed not only a higher precision in identifying relevant studies but also a substantial increase in efficiency, demonstrated by lower *L Rel* and higher *WSS* values. This indicates that the proposed approach has the potential to significantly improve efficiency and reduce costs for systematic reviews by alleviating much of the manual annotation burden from human reviewers. In addition, answer re-ranking proved to be a key success factor. This encourages advocacy for **the integration of explanatory narratives into LLMs’ responses** for future iterations. As per literature [72, 73], revealing the model’s reasoning could not only bolster transparency but also build *user trust*, which is pivotal for the adoption of automated abstract screening tools. Enabling the model to provide rationales for its answers encourages a constructive feedback loop, where user responses can inform continuous model refinement. Such an iterative process is essential for fostering a collaborative user-model relationship, ultimately enhancing the technology’s robustness and acceptability.

### Ablation Studies

Recall from Figure 2b that there are several components in our proposed framework: Question generation, question answering, question-level re-ranking and criteria-level re-ranking. Table 2 shows the results of GPT QA Soft/Hard Both ReRank, which uses all these components. This subsection investigates the impact of each component through a quasi-ablation study. We removed one or both re-ranking components, removed the question answering and question generation components, and compared the performances of the resulting methods. Thus, in addition to GPT QA Soft/Hard Both ReRank, we tested the following variants.

*•* GPT QA Soft/Hard Question ReRank: Criteria-level re-ranking component was removed. Mathematically, this is equivalent to removing *cos*(*d_i_, Q_k_*) from Eq. (3).
*•* GPT QA Soft/Hard Criteria ReRank: Question-level re-ranking component was removed. Mathematically, this is equivalent to removing *cos*(*d_i_, Q*) from Eq. (5).
*•* GPT QA Soft/Hard: Both question-level and criteria-level re-ranking components were removed.
*•* GPT Cos Sim Both: Questions were generated, but the question-answering component was removed. Both the question-level and criteria-level cosine similarities were used in scoring candidate studies. Mathematically, this means removing *score*(*d_i_, A_i,k_*) from Eq. (3).
*•* GPT Cos Sim Question: In addition to question answering, the criteria-level re-ranking component was also removed. This is equivalent to removing both *cos*(*d_i_, Q*) from Eq. (5) and *score*(*d_i_, A_i,k_*) from Eq. (3).
*•* GPT Cos Sim Criteria: The question generation component was removed, thus no questions were generated. Implicitly, the question-answering and question-level re-ranking components disappeared too. This means retreating to match each candidate study against the selection criteria. Mathematically, this is equivalent to removing (*score*(*d_i_*) from Eq. (5).

Table 3 shows the results of the ablation study. Due to space limit, we focused on three metrics: *MAP*, *R*@50% and *WSS*@95%. From the GPT QA Soft/Hard rows, we can see that our QA framework resulted in good performances, improving significantly over the non-QA baselines (first three rows) on *Intervention*, *Qualitative* and *Prognosis*. The last six rows are the performances of our framework by using either one or both re-ranking methods. Comparing them against GPT QA Soft/Hard, we see that **re-ranking was one success key** to significant performance improvement, although it is less conclusive which re-ranking method is consistently better. To gain better insights into the latter question, we averaged the performance metrics across all 31 SRs, resulting in the “Average” columns in the table. It looks that **criteria-level re-ranking maybe overall stronger than question-level re-ranking** (by comparing the GPT QA Soft/Hard Criteria ReRank rows against the GPT QA Soft/Hard Question ReRank rows). By comparing the GPT QA Soft/Hard Both ReRank rows against the four rows using one re-ranking method, we are more or less able to conclude that both re-ranking methods may have their own values in prioritising candidate studies towards facilitating systematic reviews. Meanwhile, we point out that more experiments need be done on more SRs of the *Qualitative* and *Prognostic* categories to draw more convincing conclusions on the effectiveness of the two methods of answer re-ranking, which is one of our directions for future work. Surprisingly, we found that the non-QA baselines (i.e., the GPT Cos Sim Criteria/Question/Both rows) worked extremely well on *DTA*, beating the baseline QA approaches (i.e., the GPT QA Soft/Hard rows). After analysing the qualities of generated questions for the *DTA* SRs, we guess the cause was the quality or complexity of generated questions of this SR category (more discussions in the “Quality of Question Generation” section). Nevertheless, **answering re-ranking and question answering complement each other**. This is proved by the significantly improved performances obtained by GPT QA Soft/Hard Both ReRank (i.e., the last two rows) over others (the fourth to tenth rows).

**Table 3.** Results of ablation study by removing one or several components from the processed question-answering framework.

| Models | MAP |  |  |  |  | WSS@95% |  |  |  |  |
| --- | --- | --- | --- | --- | --- | --- | --- | --- | --- | --- |
|  | Prognosis | Qualitative | DTA | Intervention | Average | Prognosis | Qualitative | DTA | Intervention | Average |
| GPT_Cos_Sim_Criteria | 0.178 | 0.082 | 0.271 | 0.315 | 0.286 | 0.239 | 0.303 | 0.600 | 0.552 | 0.536 |
| GPT_Cos_Sim_Question | 0.188 | 0.074 | 0.233 | 0.261 | 0.239 | 0.269 | 0.283 | 0.554 | 0.456 | 0.464 |
| GPT_Cos_Sim_Both | 0.191 | 0.084 | 0.261 | 0.309 | 0.278 | 0.252 | 0.310 | 0.602 | 0.544 | 0.534 |
| GPT_QA_Soft | 0.350 | 0.157 | 0.255 | 0.411 | 0.352 | 0.434 | 0.599 | 0.408 | 0.486 | 0.471 |
| GPT_QA_Hard | 0.315 | 0.110 | 0.228 | 0.356 | 0.306 | 0.417 | 0.650 | 0.364 | 0.466 | 0.450 |
| GPT_QA_Soft_Criteria_ReRank | 0.417 | 0.159 | 0.301 | 0.440 | 0.385 | 0.523 | 0.595 | 0.473 | 0.534 | 0.522 |
| GPT_QA_Hard_Criteria_ReRank | 0.417 | 0.200 | 0.310 | 0.443 | 0.392 | 0.543 | 0.608 | 0.454 | 0.532 | 0.517 |
| GPT_QA_Soft_Question_ReRank | 0.441 | 0.161 | 0.287 | 0.435 | 0.379 | 0.252 | 0.599 | 0.452 | 0.509 | 0.492 |
| GPT_QA_Hard_Question_ReRank | 0.452 | 0.156 | 0.297 | 0.435 | 0.382 | 0.501 | 0.633 | 0.441 | 0.516 | 0.503 |
| GPT_QA_Soft_Both_ReRank | 0.430 | 0.159 | 0.315 | 0.450 | 0.396 | 0.543 | 0.576 | 0.556 | 0.600 | 0.585 |
| GPT_QA_Hard_Both_ReRank | 0.429 | 0.157 | 0.322 | 0.447 | 0.395 | 0.555 | 0.601 | 0.536 | 0.592 | 0.577 |

### Quality of Question Generation

We manually checked the question qualities and found notable strengths and occasional challenges of GPT-3.5 in question generation. Figure 4a illustrates how GPT-3.5 translated a lengthy exclusion criterion into two relevant questions, *Q*_4_ and *Q*_5_. The exclusion criterion sentence (in purple and blue) was automatically reworded by GPT-3.5 (see the underlined words) in a way that a POSITIVE response consistently signifies compliance with a selection criterion. Occasionally, GPT-3.5 failed to generate completely independent questions, such as *Q*_5_ in Figure 1. This led to redundant or overlapped questions, introducing possible biases in combining answer scores. To avoid these issues, question generation may be done by humans. Less radically, it may be practical for humans to separate the selection criterion sentences and use each sentence to generate questions one by one. Humans may also manually decide how to convert a long selection criterion sentence into several questions.

**Fig. 4:**
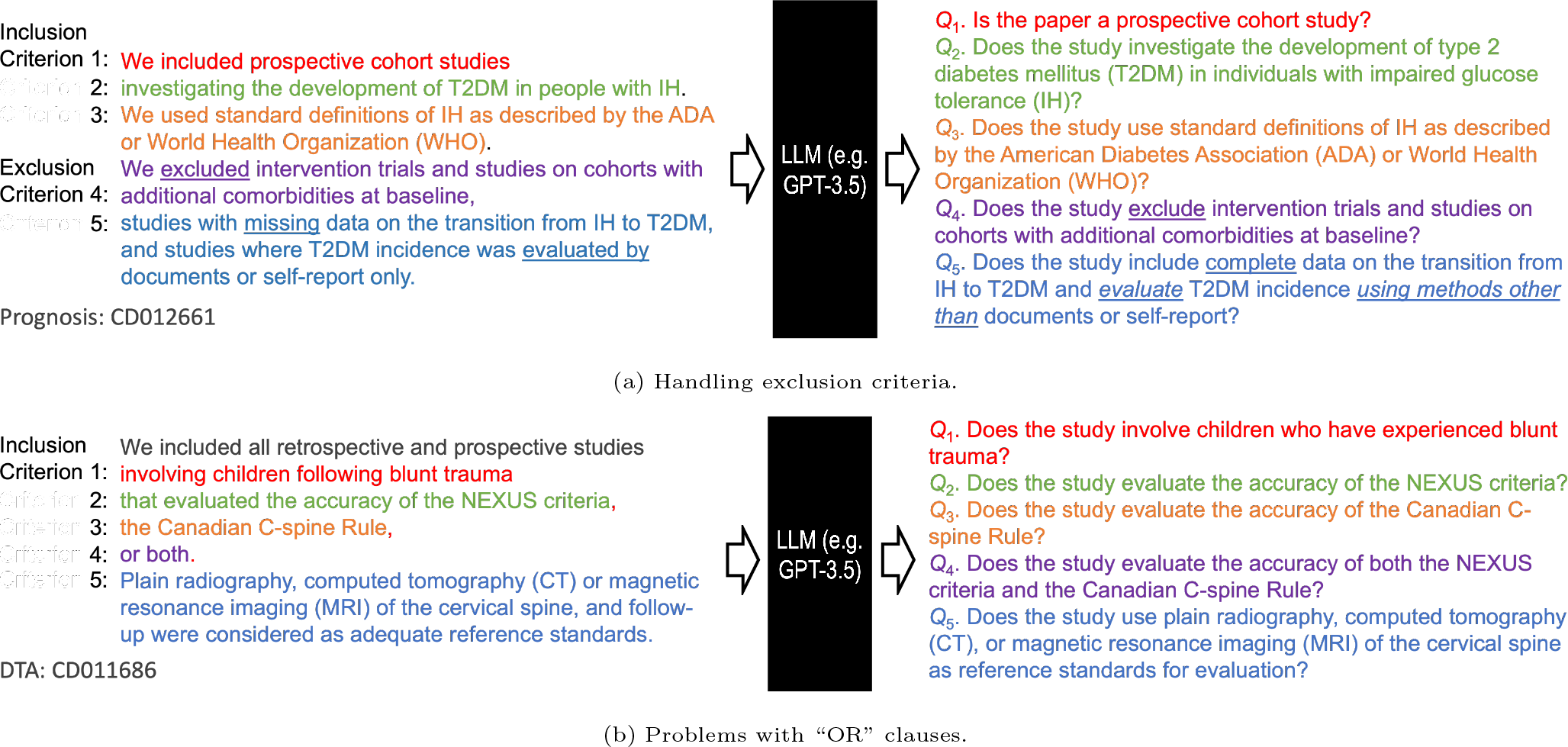
Examples of strengths and challenges of GPT-3.5 in question generation.

Notably, GPT-3.5 also displayed difficulties with complex “OR” clauses in a long selection criterion sentence, erroneously splitting them into separate questions and complicating the scoring process. In such cases, matching one question should give a POSITIVE score, but the NEGATIVE answers to other questions generated from the OR clause will mistakenly underestimate the score. Figure 4b shows an example. A single selection criterion sentence with three conditions connected by “OR” logic was split into three questions *Q*_2_ to *Q*_4_. Suppose a candidate study “evaluated the accuracy of the NEXUS criteria” and three questions are correctly answered. This results in one POSITIVE answer for *Q*_2_ (scored 1) and two NEGATIVE answers for *Q*_3_ and *Q*_4_ (scored 0). The ensemble score is 1, lower than that by rejecting answers *Q*_3_ and *Q*_4_ (two NEUTRAL answers, each scored 0.5, leading to a final score of 2), which is obviously suboptimal. This granularity of analysis points toward a need to enhance the model’s ability to discern and maintain the integrity of compound logical structures within questions. We postulate that a viable solution is to train a good question generator and analyzer to tackle these issues.

To somehow provide a quantitative evaluation of the quality of question generation, we manually annotated the “correctness” of all generated questions for all 31 SRs. The simple guidelines for scoring generated questions are as follows: 2 marks assigned to perfect question (e.g., *Q*_1_ to *Q*_4_ in Figure 1); 1 mark assigned to incomplete question, i.e., a question missing some information from the corresponding selection criterion; 1 mark assigned to each question generated from an “OR” clause (e.g., *Q*_2_ to *Q*_4_ in Figure 2b); 0 mark assigned to meaningless/redundant question (e.g., *Q*_5_ in Figure 1); and -1 mark to wrong question, i.e., a question twisted the original meaning of the corresponding criterion, which never happened in our datasets though. Table E1 in Appendix E shows the mean question scores for all four SR categories. *DTA* recorded the lowest score, 1.50, which might be the cause for the under-performance of our QA framework to our own cosine similarity baseline using selection criteria due to the reasons stated in the last paragraph. Nevertheless, the mean score is still very high, indicating no or few meaningless questions. *Prognosis* scored 2.0, meaning perfect question generation. However, it’s crucial to note that this category comprised only a single SR, potentially causing bias in evaluation. We argue that the findings underscore the possibility of delegating question generation to LLMs like GPT-3.5, though further improvement can be achieved by human intervention as a quality control measure.

### Comparing Large Language Models

Three additional successful mainstream LLMs, namely LLaMA 2, Gemini Pro and Claude 3, were compared against ChatGPT (GPT- 3.5). As shown in Table C4 in Appendix C, we found our approach performed extremely well on the *Intervention* subset. We achieved close to 100% *R*@50% for all 20 SRs in this category by at least one variant of our approach. Accordingly, because 50% samples can be safely delegated to our approach for decision, the *WSS*@95% was also very high for all *Intervention* SRs. However, GPT-3.5 struggled a bit on the *DTA* subset, mainly on two SRs (see “CD012768” and “CD012233” in Table C3 in Appendix C). Therefore, the experiments in this section focused the comparison on *DTA*. Table 4 compares the screening performances using the four LLMs as the question answering engine and two LLMs for answer re-ranking (see the “Large Language Models” section for details). For example, LLaMA QA Soft_Gemini Both ReRank means LLaMA 2 was used for question answering, and Gemini embedding was used for answer re-ranking.

**Table 4.** Performance comparison of different large language models on the DTA category.

| Models | L_Rel | MAP | R@5% | R@10% | R@20% | R@30% | R@50% | WSS@95% | WSS@100% |
| --- | --- | --- | --- | --- | --- | --- | --- | --- | --- |
| GPT_QA_Soft | 1979 | 0.255 | 0.319 | 0.495 | 0.674 | 0.765 | 0.878 | 0.408 | 0.334 |
| Llama2_QA_Soft | 1809 | 0.152 | 0.254 | 0.435 | 0.603 | 0.710 | 0.803 | 0.377 | 0.336 |
| Gemini_QA_Soft | 2487 | 0.232 | 0.309 | 0.526 | 0.678 | 0.725 | 0.847 | 0.382 | 0.235 |
| Claude_QA_Soft | 1800 | 0.163 | 0.304 | 0.410 | 0.545 | 0.763 | 0.905 | 0.436 | 0.411 |
| GPT_QA_Soft_GPT_Both_ReRank | 1136 | 0.315 | 0.438 | 0.593 | 0.766 | 0.858 | 0.941 | 0.556 | 0.506 |
| GPT_QA_Soft_Gemini_Both_ReRank | 679 | 0.347 | 0.503 | 0.669 | 0.825 | 0.886 | 0.953 | 0.630 | 0.600 |
| Llama2_QA_Soft_GPT_Both_ReRank | 1250 | 0.259 | 0.423 | 0.614 | 0.779 | 0.851 | 0.930 | 0.540 | 0.452 |
| Llama2_QA_Soft_Gemini_Both_ReRank | 833 | 0.325 | 0.514 | 0.651 | 0.825 | 0.875 | 0.951 | 0.636 | 0.540 |
| Gemini_QA_Soft_GPT_Both_ReRank | 1920 | 0.293 | 0.456 | 0.627 | 0.755 | 0.815 | 0.906 | 0.488 | 0.357 |
| Gemini_QA_Soft_Gemini_Both_ReRank | 1162 | 0.330 | 0.528 | 0.684 | 0.800 | 0.862 | 0.952 | 0.597 | 0.511 |
| Claude_QA_Soft_GPT_Both_ReRank | 1475 | 0.244 | 0.406 | 0.564 | 0.742 | 0.855 | 0.950 | 0.548 | 0.479 |
| Claude_QA_Soft_Gemini_Both_ReRank | 924 | 0.305 | 0.522 | 0.659 | 0.808 | 0.881 | 0.955 | 0.653 | 0.573 |

We have several interesting findings from Table 4. The first four rows show that, in a certain sense, Claude 3 did perform significantly better in question answering, as was expected to see. Using QA alone, it could achieve more than 90% *R*@50%. The *WSS*@95% and *WSS*@100% were both greatly improved too. We envision that **the overall effectiveness of our QA-based framework will be continuously improved with more powerful LLM-backed QA engines**. However, Gemini Pro and GPT-3.5 had better *R*@5%, *R*@10% and *R*@20% values, suggesting them to be more useful in an active learning setting. By comparing the last eight rows against the first four rows, we observe that answer re-ranking consistently improved the overall performance using different LLMs as the QA engine, further justifying the effectiveness of our proposed re-ranking methods. Additionally, Gemini embedding proved to be significantly more helpful than GPT embedding. Note that the GPT embedding model we used was “text-embedding-ada-002”, strong but much older than the Gemini embedding model “text-embedding-004”. From this angle, the results further underscore the necessity of answer re-ranking. We anticipate that **improvements in text embedding will bring added benefits to our proposed framework**. Overall, a strong QA engine (Claude 3 in our case) and a strong text embedding for re-ranking (Gemini embedding “text-embedding-004” in our case) resulted in the best-performing system, achieving a 95.5% average *R*@50% and a 0.653 average *WSS*@95%. This implies that, on average, a maximum of 65.3% of human labour can be saved with the assistance of LLMs. Finally, and not least, the performance of different LLMs varies a lot across different SRs (see Tables D1 and D2 in Appendix D). This variability provides us with an opportunity to integrate the strengths of various LLMs, which will be one of the main directions of our future work.

### Quality of Question Answering

As shown in the previous section, the quality of question answering is one key success factor for our approach. In this subsection, we attempted to evaluate the quality of responses from GPT-3.5 and three additional LLMs, including LLaMA 2 (70B), Gemini Pro and Claude 3 (see the “Large Language Models” section). We randomly selected ten samples from each systematic review in the *DT A* category. The correctness of each answer for each question on each sample was checked by the first author. To further minimise human annotation bias, we annotated these responses using three much more powerful versions of LLMs — GPT-4, Gemini 1.5 Pro, and Claude 3 Opus. The ground truth was determined by majority voting among the human annotator and all three LLM annotators. In total, 400 answers and their explanations were manually annotated, 50 per review topic.

The results in Table E2 in Appendix E show that the LLMs are nearing human-level accuracy in question answering in several instances. ChatGPT (GPT-3.5) lead with an average accuracy of 0.673 compared to 0.720 achieved by human annotators. Claude 3 Haiku and Gemini Pro also showed promising results (0.648 and 0.620, respectively). The close proximity of LLM accuracy to the human level is a promising development, particularly for applications involving a zero-shot setting, like abstract screening. This capability could be harnessed to significantly alleviate human workload by **initially filtering out straightforward cases, leaving only the most ambiguous or complex samples for human verification**. Such an approach enhances efficiency and allows human expertise to be concentrated on cases requiring more nuanced judgement, thereby optimising resource allocation and improving screening quality. Enhancing LLMs’ question-answering capability in the subject fields of systematic reviews will definitely benefit automatic abstract screening, which is one of our future research directions. In addition, it can be observed that different LLMs excelled on different review topics. Such diversity opens the gate to exploring LLM ensembles for improved accuracy.

To gain better insights, we conducted a head-to-head comparison and correlation analysis of the comparative capabilities of various LLMs. The head-to-head comparison (Figure 5a) calculated the head-to-head winning rate of each model against others, including the human annotator. As partly expected, all models outperformed LLaMA 2, with Claude 3 excelling among all LLMs but surprisingly only rivalling GPT-3.5. This corroborates with the first four rows in Table 4, where GPT-3.5 showed better performance than LLaMA 2 and Gemini across most metric while Claude 3 excelled in *L Rel*, *R*@50% and *WSS*but not others. The correlation map (Figure 5b) counted the percentage of times two LLMs gave the same answer (Positive/Neutral/Negative). Different LLMs exhibited substantial variability on the 400-question human-annotated sample set. GPT-3.5 had the highest correlation with human annotator, which corroborates its closer performance to human in the head-to-head contest. Although all models share higher answer similarity than LLaMA 2, the fact that LLaMA 2 presented good performance in term of the important metrics *L Rel* and *WSS*, which are directly linked to each other, cannot rule out LLaMA 2 from serving as an AI assistant for abstract screening, especially when open-access is factored into consideration. Nevertheless, we believe the in-depth analysis presages the synergistic potentials of different LLMs to improve the efficiency of systematic reviews.

**Fig. 5:**
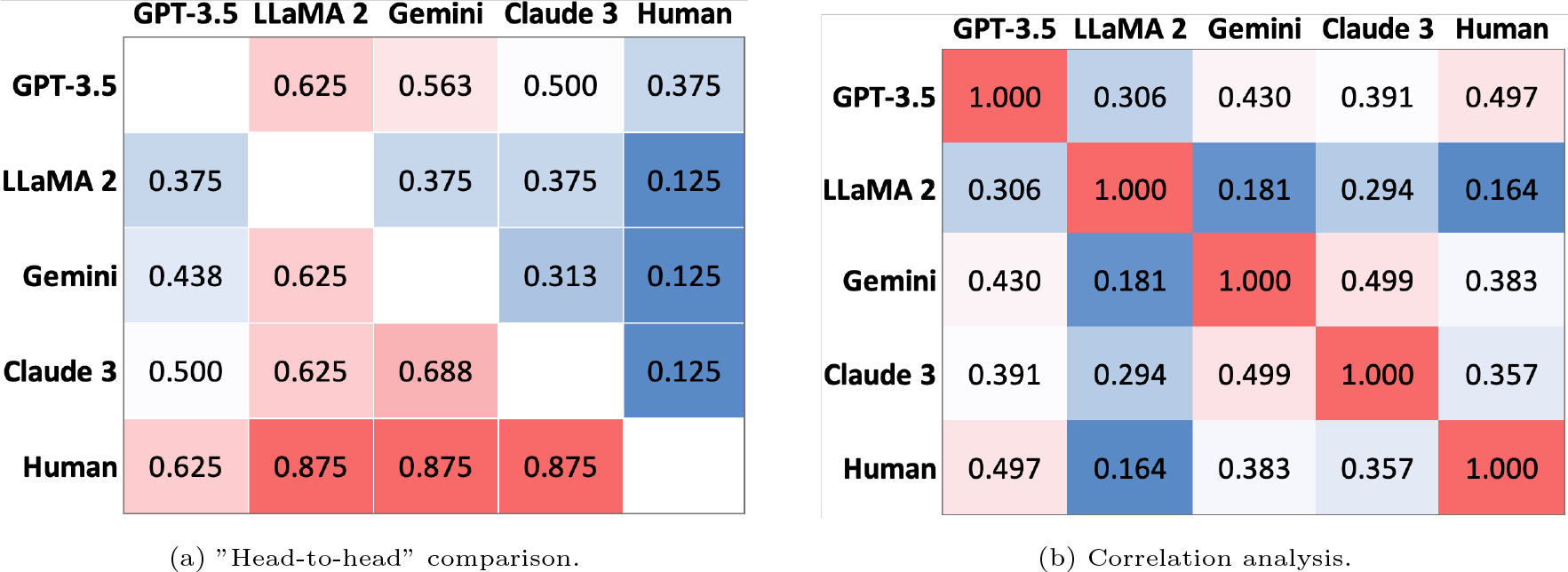
Comparisons of the question answering capabilities of different large language models.

## Conclusion

This paper proposed an effective LLM-assisted question-answering framework to facilitate abstract screening and advance automated systematic review. Our framework starts with converting selection criteria to binary questions, answering and scoring questions using LLMs, adjusting answer scores using question- and criteria-level re-rankings, and combining answer scores to prioritise candidate studies. Extensive experiments emphasised the particular pertinence of selection criteria of included studies to automated abstract screening and LLMs’ proficiency in understanding and utilising selection criteria to prioritise candidate studies. LLMs demonstrate exceptional capability in parsing and applying these criteria to discern and prioritise candidate studies to facilitate filtering relevant studies. Specifically, LLMs such as GPT-3.5 successfully handled the complexity of a mixture of inclusion and exclusion criteria by correctly phrasing the questions. The overall quality of question generation was very high based on human verification. However, it faces challenges in formulating several juxtaposed criteria with a logical OR relationship. The positive results of *L*_Rel_ (position of the last relevant study), *R*@5% (recall at top 5%), *R*@10%, *WSS*@95% (Workload Saved over Sampling at 95% recall level), and *WSS*@100% not only showed the competency of the proposed framework as a zero-shot abstract screening methodology but also indicated its potential use in reducing human effort in building a high-quality dataset for training a abstract screener. The comparative study of several mainstream LLMs in question answering, including GPT-3.5, LLaMA 2, Gemini Pro and Claude 3, has shown promising results, with some models nearing human-level accuracy. This progress suggests that LLMs can significantly reduce the human effort required in the initial filtering stage of abstract screening by handling clear-cut cases, thereby allowing human experts to focus on more ambiguous or complex instances. Such strategic deployment of LLMs enhances operational efficiency and elevates the quality of systematic reviews. Notably, the comparative analysis of various LLMs has broadened our understanding of the variability in performance across different models, highlighting the potential for leveraging the comparative strengths of diverse LLM to enhance screening accuracy. Thanks to the high *R*@95% (mostly *>* 95%), we conjecture that LLMs can replace the second reviewer by more or less safely delegating 50% screening job to LLMs or machine learning models trained with the assistance of LLMs, although large-scale validation studies are needed.

## Contributorship Statement

According to the CRediT tool, the author’s contribution is as follows. Opeoluwa Akinseloyin: Conceptualization, Methodology, Software, Validation, Investigation, Data Curation, Writing - Original Draft, Writing - Review & Editing. Xiaorui Jiang: Conceptualization, Resources, Writing - Original Draft, Visualization, Supervision, Project administration, Funding acquisition, Writing - Review & Editing. Vasile Palade: Conceptualization, Supervision, Writing - Original Draft, Writing - Review & Editing.

## Data Availability statement

The benchmark on which the experiments were done is publicly available at https://github.com/CLEF-TAR/tar/tree/master/2019-TAR/Task2. The data underlying this article are available in the article and in its online supplementary material.

